# Assessment of occupational aerosol exposure for laboratory technicians: A quantitative study using ΦX174 phage as a substitute virus

**DOI:** 10.64898/2026.06.09.26355304

**Authors:** Bo Liu, Donghua Liu, Hongwei Zhang

**Affiliations:** Department of Laboratory Medicine, Xiaogan Hospital Affiliated to Wuhan University of Science and Technology, Xiaogan, Hubei, China

## Abstract

This study aimed to clarify aerosol exposure risks throughout the workflow of a Biosafety Level 2 (BSL-2) polymerase chain reaction (PCR) laboratory, validate the suitability of the ΦX174 bacteriophage as an indicator virus, and provide evidence for biosafety control measures. The ΦX174 bacteriophage was used to simulate viral samples, and a concentration–bacteriophage plaque standard curve was constructed (*R*²=0.998). Five operational steps in a simulated PCR laboratory were quantitatively monitored for aerosol concentration using double-layer agar plates, with blank controls used to eliminate interference. Statistical analysis was employed to identify risk differences. Sample homogenization ((5.67 ± 1.23) × 10⁴ plaque-forming units (PFU)/m³) and nucleic acid extraction ((3.45 ± 0.89) × 10⁴ PFU/m³) were identified as high-/very high-risk steps. The viral load in the samples was strongly positively correlated with the aerosol concentration (r = 0.926, *P* <0.001), with aerosol levels linearly decreasing with increasing distance in high-risk steps. The ΦX174 bacteriophage demonstrated high detection sensitivity (10¹ PFU/ml) and demonstrated safety compatibility with BSL-2 laboratories. Aerosol risks in PCR laboratories exhibit step-specific differentiation, and ΦX174 serves as an ideal indicator virus. Proposed strategies such as equipment upgrades and personal protective equipment (PPE) grading can reduce exposure risks.

## Introduction

Polymerase chain reaction (PCR) technology has been widely applied in viral nucleic acid detection because of its high specificity and sensitivity. During the COVID-19 pandemic, PCR laboratories completed more than 90% of global viral diagnostic tests. However, clinical samples such as throat swabs and bronchoalveolar lavage fluid often contain pathogenic microorganisms. High-speed homogenization and pipetting operations during sample processing can aerosolize liquids into particulate matter (0.5–10 μm) that remains suspended for hours, posing risks of cross-contamination and respiratory infections [1]. Even in high-level biosafety laboratories, inadequate operational supervision may lead to aerosol leakage [2], underscoring the urgent need for aerosol control in BSL-2 PCR laboratories. Fu et al. [3] demonstrated that simplified rapid detection protocols may increase aerosol generation, highlighting the challenge of balancing efficiency and safety.

Current methods for assessing aerosol risks in PCR laboratories face two major challenges in biosafety management. First, quantitative risk assessment techniques have several limitations. Most studies rely on surface swab sampling combined with real-time quantitative PCR (qPCR) detection technology. This approach can determine only the presence of contamination but fails to accurately calculate aerosol viral concentrations or differentiate risk variations across operational steps, necessitating nontargeted protective measures [4]. Second, standardized protocols for alternative virus applications are lacking. Some studies directly use pathogenic viruses such as SARS-CoV-2, which are suitable only for BSL-3 laboratories. These studies involve higher operational complexity and safety risks, elevated costs, and potential leakage hazards, making their implementation challenging in conventional BSL-2 PCR laboratories [1]. The use of phage ΦX174 as an alternative virus requires further investigation into its biological characteristics. Orta et al. [5] elucidated the mechanism by which ΦX174 phage-encoded protein antibiotics exert their effects, whereas Parab et al. [6] demonstrated that chloramphenicol and gentamicin can reduce resistance to the ΦX174 phage by inhibiting lipopolysaccharide (LPS) mutations in *Escherichia coli*. These molecular-level studies confirm the cultivation stability and application safety of the ΦX174 phage, providing critical evidence for its suitability as an indicator virus.

Liu et al. [2] conducted a study combining numerical simulation and experimental validation in a BSL-3 laboratory and demonstrated that aerosol concentration exhibits exponential decay with increasing operational distance. Significant variations in temperature and humidity substantially affect aerosol suspension duration, providing a theoretical basis for optimizing laboratory spatial layouts. Li et al. [7] proposed enhanced personal protective measures for sample homogenization processes in COVID-19 nucleic acid testing laboratories but lacked quantified risk data, hindering the development of precise protective protocols.

With respect to disinfection efficacy, Chen et al. [8] investigated three bacteriophages (ΦX174, MS2, and Φ6) and reported differential inactivation efficiencies of gaseous ozone across different phage carriers. Notably, ΦX174 inactivation rates were significantly influenced by the surface material adsorption characteristics of phage deposition surfaces. Research by Chen’s team offers a reference for the selection of disinfection strategies following aerosol contamination in PCR laboratories and recommends the development of targeted disinfection approaches based on aerosol carrier properties.

### Indicating the status of applied research on viruses

The ΦX174 bacteriophage is an unenveloped virus that can infect *Escherichia coli* and is an ideal indicator virus for safe laboratory research. Its advantages include three aspects.

First, the cultivation efficiency is high, and clear plaques can form on double-layer agar plates within 12 to 16 h, facilitating precise counting [9]. Second, the ΦX174 phage is nonpathogenic to humans, animals, and plants, allowing operation in BSL-2 laboratories while reducing operational thresholds and costs [10]. Third, its biological characteristics, particle size (25–30 nm), surface negative charge, and aerosol behavior are highly similar to those of SARS-CoV-2 (60–140 nm) and influenza viruses (80–120 nm) [9], enabling realistic simulation of viral aerosol transmission processes. Parab et al. [6] demonstrated the stability of the ΦX174 phage cultivation system in laboratory settings by showcasing antibiotic-mediated bacterial resistance, effectively avoiding counting deviations caused by host bacterial mutations. In studies validating the application of this phage, Luo et al. [11] confirmed the efficacy of the ΦX174 phage in replacing pathogenic viruses for disinfection efficacy evaluation. Lei et al. [12] verified the quantitative detection accuracy of the ΦX174 phage at a concentration of 10^1^ PFU/ml, with a detection limit of approximately 10^2^ copies/ml, significantly outperforming traditional quantitative PCR methods. Sun et al. [10] and Lu et al. [13] emphasized the outstanding performance of ΦX174 as an indicator for aerosol transmission studies and disinfection efficacy evaluation, providing a reliable technical tool for quantitative aerosol monitoring in PCR laboratories. Additionally, Ma et al. [14] investigated the effectiveness of using the ΦX174 bacteriophage as an indicator virus for air disinfection and reported that its response to disinfectants was consistent with that of SARS-CoV-2. further validating the applicability of this method in laboratory safety research.

Although the high-risk characteristics of aerosols and the potential hazards of ΦX174 have been widely recognized, existing studies remain limited in providing comprehensive risk assessments for standard BSL-2 PCR laboratories. First, the scope of the research is relatively narrow, with a focus primarily on BSL-3 laboratories or specific PCR operational procedures (sample homogenization) and a failure to cover the complete workflow from sample collection to result analysis. Second, quantitative data on aerosol concentrations under varying viral loads and operational distances remain insufficient. Third, the technical framework is not yet well established and lacks standardized operating procedures for aerosol sampling, cultivation, and quantitative analysis. This study aims to address these research gaps by quantifying aerosol exposure risks throughout the entire BSL-2 PCR laboratory workflow using ΦX174 as a safe and effective alternative viral model.

## Methods

The lyophilized ΦX174 bacteriophage used in this experiment was purchased from the China National Institute of Metrology. Based on the validation method for the ΦX174 bacteriophage standard substance established by Liu et al. [9], professional identification confirmed that the bacteriophage was free from exogenous microbial contamination, with a purity of ≥ 99% and a concentration standard value of 10^10^ plaque-forming units (PFU)/ml, ensuring the quality, stability and quantitative accuracy of the bacteriophage during the experiment. Prior to the experiment, the lyophilized bacteriophage was reconstituted with a buffer formulation composed of a mixture of 10 mM Tris-HCl (pH 7.5) and 1 mM EDTA. This buffer was filtered through a 0.22 μm polyether sulfone filter membrane to remove bacteria, preventing exogenous contamination. The dissolution process was completed in a biosafety cabinet (Airstream® Class II Type A2, ESCO). After dissolution, the bacteriophage solution was stored at 4°C in a light-protected environment with such that that all experimental operations were completed within 24 h to avoid degradation of bacteriophage activity due to prolonged storage.

The host bacterium selected was *Escherichia coli* strain C (American Type Culture Collection, ATCC 13706), which was provided by the Microbiology Laboratory of Sichuan University. Preliminary experiments demonstrated that this strain exhibited a sensitivity to the ΦX174 bacteriophage of ≥99%, effectively supporting phage adsorption, proliferation, and plaque formation, thereby meeting the requirements for subsequent phage culture and quantitative analysis. The strain was cultured in a constant-temperature incubator (MIR-153, Panasonic Corporation) at 37°C and 180 rpm for 12 h, with cultivation termination upon reaching the logarithmic growth phase. Ultraviolet spectrophotometry measured an optical density (OD) value of approximately 0.6 at 600 nm, and plate counting confirmed a viable bacterial concentration of approximately 10^8^ CFU/mL. This stable bacterial activity characteristic renders the strain suitable for subsequent phage double-layer agar plate culture experiments.

The nutrient agar medium used in this study consisted of 10 g of peptone, 5 g of beef extract, 5 g of sodium chloride, and 15 g of agar with distilled water added to adjust the volume to 1000 mL. The pH was then adjusted to 7.2 ± 0.2 to meet the microbial growth requirements. The prepared medium was autoclaved at 121°C for 20 min to ensure sterility. After sterilization, the cooled medium at 50°C was used as the lower layer for constructing double-layer agar plates. The upper layer of the agar medium had the same composition but contained 7 g/l agar to facilitate colony formation and observation. Following sterilization and cooling to 45°C, a 1% (v/v) logarithmic-phase *Escherichia coli* suspension was added. The mixture was rapidly homogenized and immediately used to prepare the upper layer of the double-layer agar plates.

The reconstitution buffer consisted of 10 mM Tris-HCl (pH 7.5) and 1 mM EDTA. The buffer was sterilized by filtration through a 0.22 μm polyether sulfone membrane. The treated buffer was stored at 4°C and was primarily used for dissolving lyophilized ΦX174 phage powder. A 500 mg/l chlorine-containing disinfectant was mainly used for cleaning and disinfecting laboratory environments and instrument surfaces. Ethanol (75%) was primarily utilized for hand disinfection and disinfection of specific instrument surfaces to ensure aseptic conditions throughout the experimental process.

All the instruments used in the experiments received national metrological certification and were regularly calibrated and maintained to ensure their accuracy and stability. The detailed parameters of the key instruments and equipment are listed in Table 1.

**Table 1.** Parameters of key instruments and equipment.

| Instrument Name | Model | Manufacture | Core Parameters | Calibration Cycle |
| --- | --- | --- | --- | --- |
| **Biosafety cabinet** | Class II, Type A2 | ESCO (Singapore) | Downflow velocity: $0.38 \pm 0.02\ \text{m/s}$;<br>Inflow velocity: $0.57 \pm 0.03\ \text{m/s}$;<br>HEPA filtration efficiency $\geq 99.97\%$ | Once / yr. |
| **High-speed homogenizer** | Tissuelyser-48 | Shanghai Net Xin | Speed range: 1000–6000 rpm, accuracy $\pm 50$ rpm; processing capacity: 0.2–5 ml | Once / yr. |
| **PCR** | ABI 7500 | Thermo Fisher | Temperature range: $4\text{--}99^{\circ}\text{C}$, temperature | Once / 6 |
| instrument | | Scientific (America) | accuracy $\pm 0.3$ °C; Fluorescence detection channel: 5 | mo. |
| Constant temperature incubator | MIR-153 | Panasonic (Japan) | Temperature control range: 5–60 °C, temperature control accuracy $\pm 0.5$ °C; humidity control range: 30–90% RH | Once / 6 mo. |
| Colony counters | Bio-Rad Gel Doc XR+ | Bio-Rad (America) | Count range: 0–999 per plate; Count accuracy $\pm 1$; Image resolution: 5 megapixels | Once / yr. |
| Pipette | Transferpettor Research Plus | Eppendorf (Germany) | Range: 0.5–10 μL (accuracy $\pm 0.03$ μL), 10–100 μL (accuracy $\pm 0.2$ μL), 100–1000 μL (accuracy $\pm 0.5$ μL) | Once / 6 mo. |
| Laser particle counter | CLJ-BII | Beijing Sartorius | Measurement particle size range: 0.3–10 μm; sampling flow rate: 2.83 l/min; counting accuracy $\pm 10\%$ | Once / yr. |

### Study protocol design

The specific steps to construct a concentration‒plate number standard curve for the ΦX174 bacteriophage are as follows: (1) Freeze-dried ΦX174 phage powder was dissolved in reconstitution buffer to prepare a stock solution with a concentration of 10^10^ PFU/ml. Serial dilutions were subsequently performed using a 10-fold gradient dilution method to obtain phage solutions with concentrations of 10^1^, 10^2^, 10³, 10⁵, 10⁷, 10⁹, and 10¹⁰ PFU/ml. To minimize the impact of random errors on the experimental results, five parallel samples were used for each concentration gradient. (2) To prepare double-layer Petri dishes, 20 ml of cooled nutrient agar medium (50°C) was slowly poured into sterile 90 mm Petri dishes. The dishes were placed horizontally and allowed to solidify completely to form a base layer, after which 5 mL of cooled upper agar medium containing a 1% *Escherichia coli* suspension was rapidly added. Gentle shaking was performed to ensure a uniform distribution of the upper medium.

After solidification, the double-layer agar plates were incubated at room temperature for 30 min to achieve complete stabilization for subsequent experiments. (3) During phage coating and incubation, 100 μL of each phage solution concentration was accurately pipetted onto the surface of double-layer agar plates using sterile pipettes (Transferpettor Research Plus, Eppendorf). The solution was evenly spread across the plate surface using a sterile L-shaped glass spreader. The plates were left to stand for 5 min to ensure complete agar absorption and were then inverted and incubated in a 37°C constant temperature incubator for 12–16 h until transparent bacteriophage plaques (diameter of 1–2 mm, with neat edges and no turbidity) formed on the surface. (4) During standard curve construction, PFU were counted on each plate using a colony counter (Bio-Rad Gel Doc XR+, Bio-Rad), and the average plaque count at different concentrations was calculated. The logarithm of the phage concentration (lg PFU/ml) was plotted on the x-axis, with the corresponding plaque count (PFU) on the y-axis. Linear regression analysis was performed using Origin 2023 software (OriginLab Corporation) to determine the regression equation and correlation coefficient (*R*²); to ensure method reliability, the correlation coefficient *R*² must be ≥0.99. The results of plaque counts for different concentrations of the ΦX174 phage in preliminary experiments are shown in Table 2.

**Table 2.** Number of phage spots corresponding to ΦX174 phages at different concentration gradients.

| $\Phi$ X174<br>concentration<br>(PFU/ml) | $10^1$ | $10^2$ | $10^4$ | $10^6$ | $10^8$ | $10^{10}$ |
| --- | --- | --- | --- | --- | --- | --- |
| Mean number of<br>plaques (PFU) | $1.2 \pm 0.3$ | $11.5 \pm 1.2$ | $1025.1 \pm 23.4$ | $10234.8 \pm 156.9$ | $101234.7 \pm 1023.6$ | $1002345.6 \pm 8901.2$ |
| Standard Deviation<br>(SD) | 0.3 | 1.2 | 23.4 | 156.9 | 1023.6 | 8901.2 |
| $R^2$ price | - | - | - | - | - | 0.998 |

A standard curve between the ΦX174 phage concentration and plaque count was constructed. The linear regression equation was y = 1.05x + 0.42 (where y represents phage quantity and x denotes logarithmic units of phage concentration: lg PFU/ml). The correlation coefficient *R*² was 0.998, and residual analysis revealed that all the residuals were within the ±0.5 range, indicating no significant systematic error. These results demonstrate that the quantitative method exhibits excellent linearity and can be utilized for subsequent aerosol concentration calculations.

### Aerosol monitoring

Aerosol monitoring in PCR laboratories was conducted according to the following procedures: (1) Preparation of simulated samples: On the basis of the actual viral load of clinical PCR laboratory samples, five concentration gradients of ΦX174 phage simulated samples were prepared, with concentration gradients of 10³, 10⁵, 10⁷, 10⁹, and 10¹⁰ PFU/ml. Twenty parallel samples were prepared for each concentration gradient, with each sample volume of 3 ml matching the storage volume of clinical throat swab specimens to ensure experimental reliability. (2) Simulation of operational procedures: Five experienced technicians, each with over three years of PCR operation expertise, carried out full-process simulated PCR laboratory operations within a biosafety cabinet (Airstream® Class II Type A2, ESCO). To ensure the reliability and reproducibility of the results, they followed standardized operating procedures. Each step of the operation was meticulously repeated ten times, effectively reducing the potential variability in experimental results that could be introduced by individual operator differences.

The detailed operational procedures and aerosol collection steps are described as follows. During the sample collection phase, a sterile throat swab was immersed in 3 mL of simulated sample, followed by gentle insertion into a sampling tube containing 3 mL of preservation solution. The swab was rotated and pressed five times to ensure complete dissolution of the sample in the preservation solution. After discarding the swab, the sampling tube cap was sealed. This process took 5 min. In the sample homogenization phase, the sampling tube was placed into the adapter of a high-speed homogenizer (Tissuelyser-48, Shanghai Net Xin). The homogenizer speed was 4000 rpm, and the homogenization time was 45 s on the basis of the sample characteristics. The homogenization process was initiated and completed within 2 min. During the nucleic acid extraction phase, a magnetic bead nucleic acid extraction kit (TianGen Bio-Technology, DP302, Beijing, China) was used at the nucleic acid extraction workstation. First, 200 μL of the homogenate sample was transferred into reaction wells using a sterile pipette (Transferpettor Research Plus, Eppendorf). The samples were then lysed for 10 min at 56°C in a constant-temperature incubator (MIR-153, Panasonic Corporation) to fully release the nucleic acids. Impurities were removed through three consecutive washes, followed by elution with 50 μL of elution buffer for 15 min. During the PCR amplification phase, 20 μL of elution buffer was mixed with 30 μL of the PCR mixture and transferred to PCR tubes. The tubes were sealed and loaded into a PCR instrument (ABI 7500, Thermo Fisher Scientific). The amplification program was as follows: 5 min of predenaturation at 95°C, followed by 40 cycles and an additional 10-min extension at 72°C, with the entire process lasting 120 min. To analyze the results, sequence detection software on the PCR instrument was used to analyze amplification curves to determine sample positivity, and the results were recorded. This step took 5 min to complete. Each operational stage involved the placement of double-layer agar plates at critical locations according to the airflow distribution pattern of the work area (predetermined by a laser particle counter [CLJ-BII, Sartorius]) to ensure comprehensive and accurate capture of aerosols generated during each phase. The detailed sampling protocols are presented in Table 3. After sampling was completed, the collected plates were meticulously labeled with operational steps, sample concentration, sampling distance, operator information, and collection time for subsequent data organization and analysis. The plates were inverted and incubated in a 37°C constant temperature incubator (MIR-153, Panasonic Corporation) for 12 to 16 h until bacteriophage plaque formation occurred. The number of plaques on each plate was counted, and the aerosol concentration (PFU/m³) was calculated using a standard curve established through preliminary

**Table 3.** Main experimental sampling scheme.

| Operation Steps | Sampling Location | Sampling Distance<br>(distance from the operation center) | Number of tablets<br>(per visit) | Duration of sampling |
| --- | --- | --- | --- | --- |
| **Sample Collection** | The sampling tube is evenly distributed around 360° | 10 cm, 20 cm, 30 cm | 6<br>(Each distance 2) | Leave it standing for 30 min after operation |
| **Sample Homogenization** | The homogenizer is evenly distributed around 360° | 10 cm, 20 cm, 30 cm, 40 cm, 50 cm | 10<br>(2 distance each) | Leave it standing for 30 min after operation |
| **Nucleic Acid Extraction** | Extract the workbench surface | 10 cm, 20 cm, 30 cm | 6<br>(Each distance 2) | Leave it standing for 30 min after operation |
| **PCR Amplification** | In front of the inlet of the PCR instrument | 10 cm, 20 cm | 4<br>(Each distance 2) | After the reactor is placed, it is left standing for 30 min |
| **Interpretation of Results** | Around the computer console | 10 cm | 2 | Keep it on during operation |

The aerosol concentration was determined using the following equation:

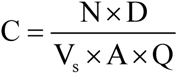 where C is the aerosol concentration (PFU/m³), N is the plaque count, D is the dilution factor, V_s_ is the spread volume (l), A is the effective sampling area of the plate (m²), and Q is the volume of air passing through the sampling area (m³). In this study, no additional dilution was performed ( D = 1), and a fixed spread volume of 0.1 ml (1×10^-4^l) was used. With a plate diameter of 90 mm, the sampling area was 0.00636 m². The air volume Q was derived from the downward airflow velocity of 0.38 m/s inside the biosafety cabinet, multiplied by the sampling duration of 30 min (1800 s), yielding Q = 0.00636m^2^ × 0.38m / s × 1800s ≈ 4.32m^3^ .

### Blank control experiment to exclude environmental interference

(1) Blank Sample Preparation: The volume and preservation conditions of the blank samples were identical to those of the simulated samples. A total of 20 blank samples were prepared using resuspension buffer devoid of ΦX174 bacteriophage as control samples. (2) Blank Sample Processing and Sampling: The same technical team strictly followed the experimental protocol for blank sample operations. At each step, double-layer agar plates identical in quantity to those used in the main experiment were employed for sampling, with the cultivation conditions maintained consistent with those of the primary experiment. (3) Interference Identification Criteria: After cultivation was completed, researchers observed all the blank control plates. The absence of bacterial plaques on all the control plates confirmed the absence of interfering microorganisms capable of lysing *Escherichia coli* in the experimental environment, thereby ensuring reliable primary experimental data. The presence of bacterial plaques indicated environmental interference requiring immediate termination of the experiment. In such cases, researchers thoroughly disinfected biosafety cabinet walls and instrument surfaces using a 500 mg/l chlorine disinfectant solution. All culture media were replaced and subjected to secondary sterilization until no bacterial plaques remained in the blank control experiments before proceeding with the main experiment.

### Statistical methods

Statistical analysis was conducted using SPSS 26.0 software (IBM Corp., Armonk). Since the aerosol concentration data usually follow a skewed distribution, the concentration values were first transformed using the common logarithm (lg PFU/m³). The Shapiro-Wilk test was used to verify the normality of the transformed data (*P* > 0.05), and the Levene test was used to assess the homogeneity of variance. After logarithmic transformation, the aerosol concentration data at each stage were normally distributed and met the requirement of homogeneity of variance, so parametric tests were adopted. The mean (x̄), standard deviation (SD), median (M), and 95% confidence interval (95% CI) of aerosol concentration at each stage were calculated, and these statistical indicators were used to describe the data distribution and intuitively reflect the overall situation of aerosol concentration during the operation stages. One-way ANOVA was used to compare the differences in aerosol concentration among different operation stages, sample concentrations, and sampling distances. When statistical significance was shown in the analysis (*P* < 0.05), the Tukey method was applied for multiple comparisons to identify the specific groups with significant differences. Pearson correlation analysis was used to test the relationship between sample concentration (lg PFU/m³) and aerosol concentration (lg PFU/m³), and the correlation coefficient (r) was calculated.

For the identified high-risk operations (sample homogenization and nucleic acid extraction), a linear regression analysis was conducted with sampling distance as the independent variable and aerosol logarithmic concentration as the dependent variable. A concentration-distance regression equation was established to determine the spatial distribution pattern of aerosol concentration in key operation stages.

### Quality control measures

In this study, rigorous quality control measures were implemented across five dimensions to ensure the scientific validity, reproducibility, and accuracy of the research findings: (1) Laboratory personnel received standardized professional training covering operational protocols, equipment operation methods, and biosafety procedures. Participants underwent mandatory competency assessments prior to experimental participation. Strict adherence to standardized protocols was maintained to prevent data bias caused by individual operational variations. (2) Key instruments, including biosafety cabinets, homogenizers, and PCR systems, were comprehensively calibrated with the parameters recorded to meet the experimental requirements. Real-time airflow velocity monitoring within biosafety cabinets maintained operating conditions within the range of 0.38 ± 0.02 m/s, ensuring smooth experimental execution. (3) After the ΦX174 phage mock samples were prepared, 10% of the samples were randomly selected for qPCR concentration validation using the primers F-5’-GCTGTTGTTGCTGTTGCTGT-3’ and R-5’-CAGCAGCAGCAGCAGCAGTA-3’.

Concentration variations were controlled within 5% to ensure accuracy, providing reliable baseline data for subsequent aerosol concentration quantification. (4) Prepared double-layer agar plates underwent 5% sterility testing. The Petri dishes were incubated in 37°C incubators for 24 h to observe bacterial growth, ensuring 100% sterility rates and preventing contamination from affecting the experimental results. (5) The Grubbs test (α = 0.05) was employed to screen for outliers in the experimental data. If outliers were detected (*P* < 0.05), the experiment was repeated to ensure data reliability and avoid interference with the analytical results and conclusion derivation.

## Results

In preliminary experiments, ΦX174 phage solutions treated with a concentration gradient of 10¹–10¹⁰ PFU/ml formed distinct and well-defined phage plaques on double-layer agar plates, with no observed phage plaque fusion or microbial contamination. These results indicate optimal phage culture conditions and that sterility was maintained throughout the experimental process. Statistical analysis of phage plaque counts across different concentration gradients revealed a significant linear growth trend with increasing phage concentration. The linear regression equation established between the logarithmic concentration (lg PFU/ml) and the phage plaque count (PFU) was y = 1.05x + 0.42 (where y represents the phage plaque count and x represents the logarithm of the phage concentration). The correlation coefficient *R*² reached 0.998, indicating highly statistically significant results.

Residual analysis of the standard curve revealed that all the residuals were within the range of ±0.5, with no significant systematic errors detected. These results indicate that the quantitative method exhibits excellent linear characteristics and accurately reflects the relationship between the ΦX174 phage concentration and the number of plaques formed, making this method suitable for subsequent calculations of aerosol concentrations in primary experiments.

The blank control experiment played a critical role in validating the reliability of the primary experimental data. No bacteriophages were observed on the double-layer agar plates of any of the blank control groups after incubation, and the results of the sterility test confirmed the absence of microbial growth. These findings indicate that the experimental environment (including air within the biosafety cabinet, instrument surfaces, and culture media) contained no interfering microorganisms capable of lysing *Escherichia coli*. All observed bacteriophages in the main experiment originated from aerosols of the ΦX174 bacteriophage generated during the operational procedures, with no interference from environmental factors affecting the experimental data. Therefore, the primary experimental data demonstrate reliability and are suitable for subsequent analysis and conclusion derivation.

### Main experimental results

Statistical analysis of the core experimental data revealed significant differences in aerosol concentration across the five PCR laboratory operational procedures (*F* = 156.32, *P* < 0.001). The highest concentration measured during homogenization was (5.67 ± 1.23) × 10⁴ PFU/m³, which was 164 times greater than the concentration measured during sample collection and 64.7 times greater than that measured during result analysis. These data indicate that the homogenization stage is the primary source of aerosol contamination in PCR laboratories, posing significant risks. The nucleic acid extraction procedure followed closely, with an average concentration of (3.45 ± 0.89) × 10⁴ PFU/m³, and was also classified as a high-risk operational procedure. In contrast, the aerosol concentrations generated during sample collection, PCR amplification, and result analysis were relatively low (all below 2×10³ PFU/m³), indicating lower risk levels. Detailed statistical data on the aerosol concentrations at each operational stage are presented in Table 4.

**Table 4.** Aerosol concentration data at each stage.

| Operation Steps | Sample concentration range (PFU/ml) | Mean aerosol concentration (PFU/m<sup>3</sup>) | Standard Deviation (SD) | Median (M) | 95% Confidence Interval | The multiple relationship with the homogenization of the sample |
| --- | --- | --- | --- | --- | --- | --- |
| Sample Collection | 10<sup>3</sup>-10<sup>10</sup> | $(3.45 \pm 0.12) \times 10^2$ | $0.12 \times 10^2$ | $3.42 \times 10^2$ | $(3.38 - 3.52) \times 10^2$ | 1/164 |
| Sample Homogenization | 10<sup>3</sup>-10<sup>10</sup> | $(5.67 \pm 1.23) \times 10^4$ | $1.23 \times 10^4$ | $5.58 \times 10^4$ | $(5.32 - 6.02) \times 10^4$ | 1 |
| Nucleic Acid Extraction | 10<sup>3</sup>-10<sup>10</sup> | $(3.45 \pm 0.89) \times 10^4$ | $0.89 \times 10^4$ | $3.39 \times 10^4$ | $(3.21 - 3.69) \times 10^4$ | 1/1.64 |
| PCR Amplification | 10<sup>3</sup>-10<sup>10</sup> | $(1.23 \pm 0.34) \times 10^3$ | $0.34 \times 10^3$ | $1.18 \times 10^3$ | $(1.12 - 1.34) \times 10^3$ | 1/46.1 |
| Interpretation of Results | 10<sup>3</sup>-10<sup>10</sup> | $(8.76 \pm 0.21) \times 10^2$ | $0.21 \times 10^2$ | $8.72 \times 10^2$ | $(8.61 - 8.91) \times 10^2$ | 1/64.7 |

To further clarify the differences in aerosol concentration across various operational stages, this study employed the Tukey method for multiple comparison analysis. The results demonstrated statistically significant differences in aerosol concentration during the homogenization stage compared with the other four stages (*P* < 0.001), indicating a significantly higher aerosol pollution risk during this stage. The nucleic acid extraction stage also significantly differed from the sample collection, PCR amplification, and result analysis stages (*P* < 0.001), suggesting that the aerosol pollution risk was markedly greater for the nucleic acid extraction stage than for the other three low-risk procedures. No statistically significant differences were detected between the sample collection stage and the PCR amplification stage (*P* = 0.068) or between the PCR amplification stage and the results analysis stage (*P* = 0.082). Notably, the aerosol concentration during the sample collection stage in laboratory operations was significantly lower than that during the result analysis stage (*P* = 0.032).

### Correlation between viral load and aerosol concentration in the sample

The results of this study revealed a significant positive correlation between the simulated sample concentration and aerosol concentration (r = 0.926, *P* < 0.001), indicating that higher viral loads in samples correlate with elevated aerosol concentrations during operations and consequently increased aerosol contamination risk. The correlation patterns varied across operational stages, with homogenization having the strongest correlation (r = 0.941, *P* < 0.001), suggesting that the viral load in this stage most significantly influenced the aerosol concentration. The nucleic acid extraction stage showed a moderate correlation, whereas the sample collection, PCR amplification, and result analysis stages exhibited weaker correlations, indicating relatively minor effects of the viral load on aerosol concentrations in these low-risk procedures.

Taking the homogenization stage as an example, when the simulated sample concentration increased from 10³ PFU/ml to 10¹⁰ PFU/ml, the aerosol concentration increased from (1.23 ± 0.15) × 10³ PFU/m³ to (8.98 ± 1.56) × 10⁴ PFU/m³, indicating an approximately linear growth trend. Linear regression analysis of sample concentration versus aerosol concentration data during this phase yielded the regression equation y = 0.89x + 1.23 (where y represents the logarithmic unit of aerosol concentration: lg PFU/m³, and x represents the logarithmic unit of sample concentration: lg PFU/ml), with a correlation coefficient *R*² = 0.857, indicating that the equation effectively fits the relationship between sample concentration and aerosol concentration. Aerosol concentration data obtained during homogenization processes at different sample concentrations are detailed in Table 5.

**Table 5.** Aerosol concentration data in homogenization step at different sample concentrations.

| Sample concentration (lg PFU/ml) | 3 | 5 | 7 | 9 | 10 |
| --- | --- | --- | --- | --- | --- |
| Mean aerosol concentration (lg PFU/m<sup>3</sup>) | 3.09 ± 0.05 | 3.82 ± 0.11 | 4.56 ± 0.15 | 5.21 ± 0.18 | 5.95 ± 0.22 |
| Standard Deviation (SD) | 0.05 | 0.11 | 0.15 | 0.18 | 0.22 |
| Correlation Coefficient (r) | - | - | - | - | 0.941 |
| $R^2$ price | - | - | - | - | 0.857 |

### Spatial distribution characteristics of aerosols in high-risk step zones

For the two high-risk operational steps of homogenization and nucleic acid extraction with the highest aerosol concentrations in the samples, spatial distribution patterns of aerosol concentrations were analyzed to provide data support for optimizing laboratory equipment layout and determining safe operating distances for personnel.

During the sample homogenization phase, linear regression analysis revealed a significant linear decline in aerosol concentration with increasing sampling distance, with the regression equation being y = -0.032x + 60.21 (where y represents the logarithmically transformed aerosol concentration in units of lg PFU/m³ and x denotes the sampling distance in centimeters). The aerosol concentration peaked at 10 cm from the operation center (8.76 ± 1.89) × 10⁴ PFU/m³, but decreased to (1.02 ± 0.34) × 10⁴ PFU/m³ at 50 cm, representing only 11.6% of the concentration at 10 cm. These results unequivocally demonstrate the significant impact of distance on aerosol concentration.

During the nucleic acid extraction phase, the aerosol concentration also exhibited distance-dependent attenuation characteristics, with its linear regression equation being y = - 0.028x + 5.87 (where y represents the logarithmically transformed aerosol concentration in pgF/μm² and x denotes the sampling distance in cm). At the 10 cm sampling point, the aerosol concentration reached (5.43 ± 1.01) × 10⁴ pgF/μm²; by the 50 cm sampling point, the concentration decreased to (0.98 ± 0.21) × 10⁴ pgF/μm², representing only 18.0% of the concentration at the 10 cm point. Table 6 presents detailed aerosol concentration data obtained at various sampling distances during the two high-risk procedures.

**Table 6.**
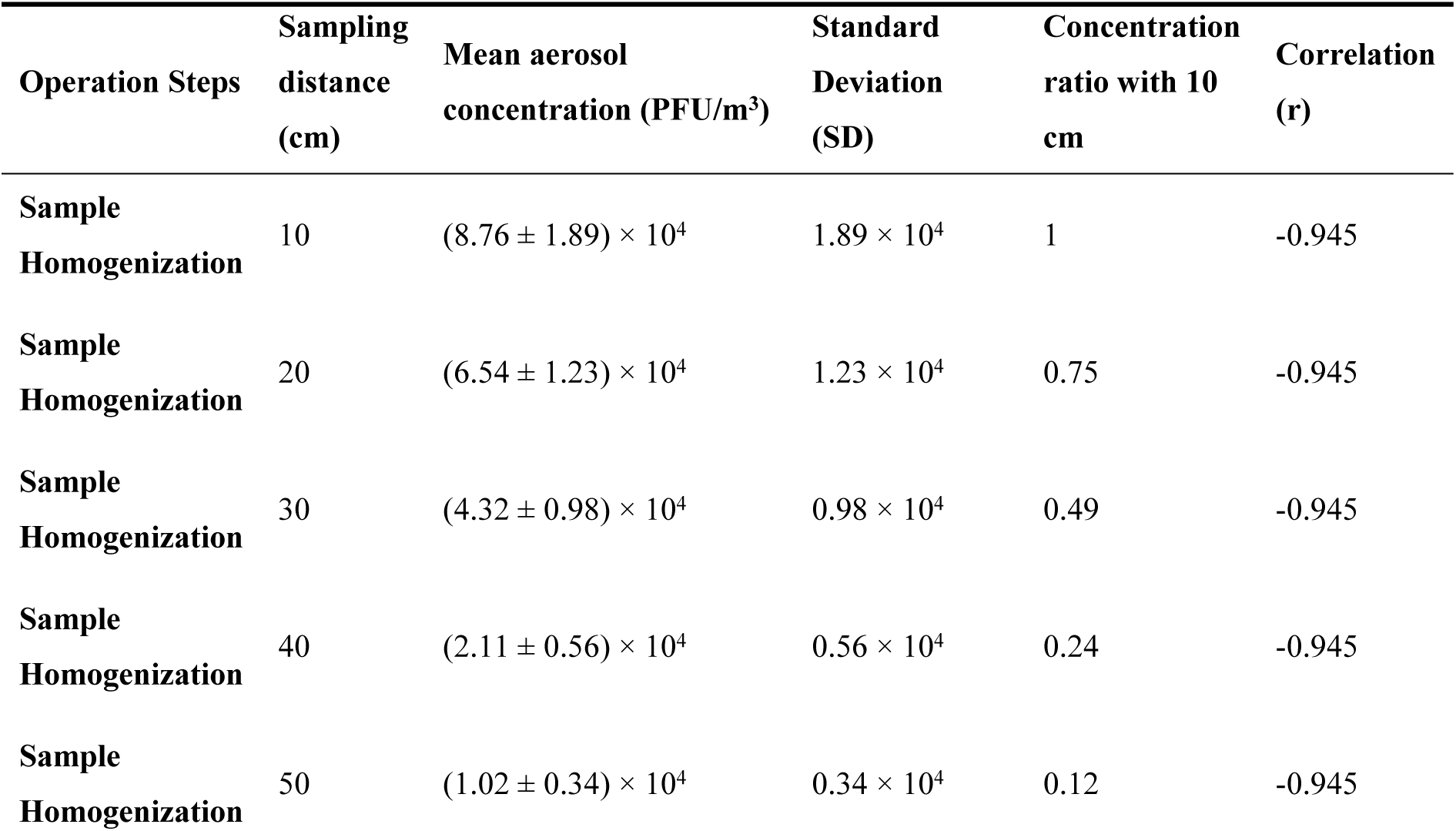
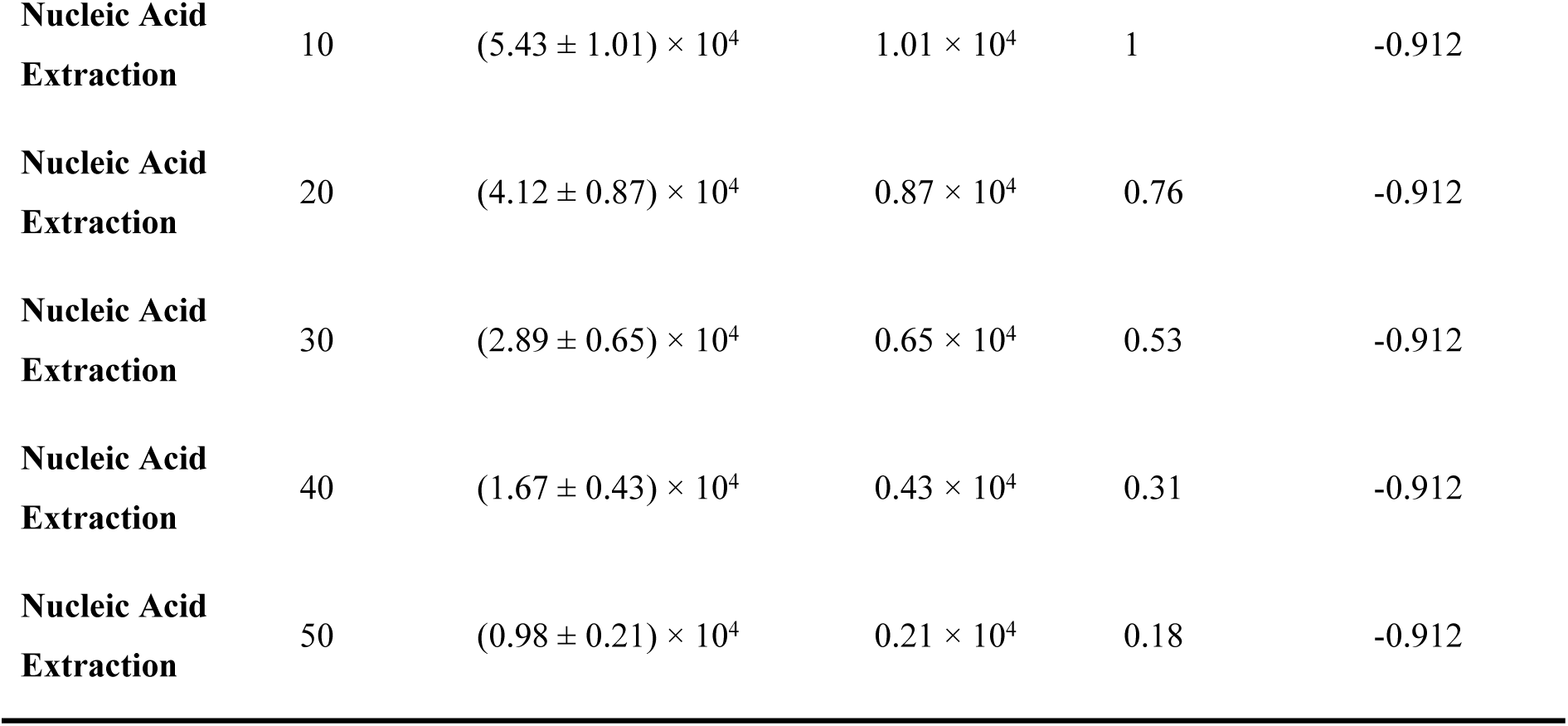
Detailed data of aerosol concentration at different sampling distances in high-risk links.

### Differences in aerosol concentration among different operators

In this study, a statistical analysis of aerosol concentrations generated by five operators at different operational stages was conducted to investigate the impact of operator proficiency on aerosol concentration. Although differences in aerosol concentrations were observed among the five operators, none of these differences were statistically significant (*F* = 2.13, *P* = 0.086). Specifically, operators with higher proficiency (those with ≥5 years of PCR testing experience) produced an average aerosol concentration of (3.21 ± 0.78) × 10⁴ PFU/m³, which was slightly lower than that of operators with lower proficiency (those with 1–3 years of PCR testing experience, averaging (3.89 ± 0.92) × 10⁴ PFU/m³). However, this difference remained not statistically significant (*P* = 0.125). These findings indicate that operator proficiency has a minimal effect on aerosol concentration and that risk variations are determined primarily by the mechanical characteristics of operational procedures, such as high-speed rotation of homogenizers and pipetting actions, rather than individual operator differences. Therefore, in PCR laboratory biosafety management, greater emphasis should be placed on optimizing operational procedures and improving equipment performance to reduce aerosol contamination risks rather than relying solely on operator proficiency.

## Discussion

### Aerosol generation mechanisms and risk level stratification analysis for each operational procedure

On the basis of quantitatively measured aerosol concentration data, risk levels for various operational procedures in PCR laboratories were classified. The sample homogenization step was categorized as an extremely high-risk procedure because of the generation of ultra-high aerosol concentrations from high-speed mechanical oscillation. The nucleic acid extraction step was classified as high risk, primarily stemming from aerosol production during pipetting, mixing, and centrifugation processes. Sample collection, PCR amplification, and result analysis procedures were classified as low-risk operations, as they involve minimal liquid handling or are performed within closed systems.

### High-risk step: nucleic acid extraction

Aerosol generation during nucleic acid extraction primarily originates from three critical operational steps. First, during pipetting procedures, rapid aspiration creates negative pressure within the pipette tip, leading to solution atomization. Meanwhile, during the ejection phase, direct activation of the second setting causes the residual solution to be forcefully ejected, particularly in filter-free pipettes, and aerosols may penetrate equipment, resulting in contamination and cross-contamination [4]. Second, during reagent mixing, vigorous shaking during lysis buffer–sample mixing generates excessive foam, releasing aerosol particles ranging from 0.1–1 μm in size, which exhibit enhanced suspended diffusion ability. Third, during centrifugation operations, inadequate sealing of centrifuge tubes allows high-speed rotation (12000 rpm) to induce solution leakage, while aerosols released upon container opening rapidly disperse into the environment. Given these risks, this step is classified as a "high-risk" procedure requiring enhanced operational training and protective equipment.

### Low-risk operational steps: sample collection, PCR amplification, and result analysis

Throughout the entire sample collection process, the sampling tube remained fully sealed except for brief openings during pharyngeal swab insertion. The operators performed manual maneuvers with minimal mechanical force to prevent solution atomization while maintaining low aerosol concentrations. PCR amplification was conducted in sealed PCR instruments, with reaction tubes isolated from ambient air through hermetic lids or optical films. Although trace aerosols may be generated during pipetting procedures, the short operation duration and stable airflow ensure negligible environmental and personnel exposure. During the analysis of the results, amplification curve data were processed exclusively via software to avoid sample contact with the solutions. Residual aerosols were diluted through biosafety cabinets and laboratory ventilation filtration systems, reducing concentrations to safe levels. Consequently, all three operational phases were classified as "low-risk" procedures requiring only standard protective measures to ensure safety.

### Scientific basis and advantage of phage ΦX174 as an indicator virus

#### Biological characteristics are highly consistent with pathogenicity

The bacteriophage ΦX174 is highly biocompatible with clinically prevalent pathogenic viruses such as SARS-CoV-2 and influenza viruses. With a particle size range of 25–30 nm, its dimensions overlap with those of SARS-CoV-2 (60–140 nm) and influenza (80–120 nm) viruses, enabling precise simulation of pathogenic viral behavior during aerosolization. All three phages possess negatively charged surfaces, with ΦX174 demonstrating a diffusion coefficient of approximately 1.5×10⁻⁵ m²/s—comparable to that of SARS-CoV-2 (1.2×10⁻⁵ m²/s)—ensuring a consistent dispersion velocity and range in air.

Additionally, ΦX174 exhibits virus-like tolerance, maintaining viability for 24 h at 25°C and 50% relative humidity. This authentic replication of viral activity and transmission processes in PCR laboratories effectively mitigates misjudgments caused by variations in indicator virus characteristics.

### The advantages of quantitative detection technology are significant

Compared with traditional qPCR methods, the dual-layer agar plate culture technique utilizing ΦX174 bacteriophages has significant advantages in terms of quantitative detection. This approach achieves a detection limit as low as 10^1^ PFU/ml, surpassing the qPCR upper limit of 10^2^ copies/ml, thus enabling precise quantification of low-concentration aerosols while effectively capturing trace laboratory contaminants. The cultured phage plaques exhibit a regular morphology with well-defined boundaries, yielding counting errors ≤5%. Unlike qPCR methods that are dependent on standard calibrators, this technology avoids quantitative deviations caused by variations in primer efficiency or interference from inhibitors. Cost-effectively, each sample costs approximately 5 CNY (about 1/5 of qPCR’s 25 CNY per sample), making it particularly suitable for high-frequency aerosol monitoring scenarios in laboratories.

### The safety compliance meets the requirements of a BSL-2 laboratory

The safety characteristics of the ΦX174 phage ensure its suitability for conventional BSL-2 standard PCR laboratories. This phage exhibits strict host specificity, infecting only *Escherichia coli* without causing pathogenic effects on humans, animals, or plants, posing no health risks to operators or environmental hazards. The experimental process requires neither the positive pressure protective suits used in BSL-3 laboratories nor dedicated ventilation systems, enabling complete operational procedures within standard BSL-2 facilities, thereby significantly reducing operational thresholds and costs. Additionally, as this phage lacks gene integration capability, genomic recombination with host bacteria is effectively prevented, completely eliminating biosafety risks. The overall safety characteristics of ΦX174 are in full accordance with the fundamental principles governing the use of non-pathogenic or low-risk microorganisms under standard BSL-2 containment protocols.

### Optimization strategies for PCR laboratory biosafety based on the findings of this study

#### High-risk operation protocols and equipment upgrades

(1) Sample homogenization treatment: Regarding equipment selection, the use of a sealed negative pressure homogenizer, such as the KAT25 CNC ultra-high-speed homogenizer (KAT25 digital ULTRA-TURRAX®, IKA), is recommended. This device is equipped with an integrated HEPA filter that can directly filter and discharge aerosols generated during homogenization. Studies have shown that such equipment can reduce aerosol concentrations by 70–80% [7]. For parameter optimization of pharyngeal swab samples, the homogenizer rotation speed should be set to 3000–3500 rpm, and the homogenization time should be controlled within 30–45 s to avoid excessive homogenization leading to excessive aerosol generation. Operational protocols require (a) always inspecting the sampling tube cap for seal integrity to prevent loosening before homogenization and (b) waiting for complete equipment shutdown (approximately 30 s) after homogenization before opening the cap to prevent sudden leakage of high-pressure aerosols from the tube. (2) Nucleic acid extraction: The use of Eppendorf filter tips equipped with built-in filters (Eppendorf Filter Tips, Eppendorf) is mandatory to intercept aerosols in pipettes, thereby preventing equipment contamination. A MagNA Pure 96 fully automated sealed nucleic acid extraction system (Roche MagNA Pure 96, Roche) is recommended. This system reduces aerosol leakage risk by more than 90% when the entire procedure is completed within a sealed chamber. During operational training, regular pipetting drills should emphasize the "slow aspiration and slow dispensing" technique: aspirate slowly by positioning the pipette tip 1–2 mm below the liquid surface and dispense by pausing for 2 s at the first stop before a second stop to minimize aerosol generation. For reagent optimization, a low-foaming lysis buffer containing 0.1% defoamer should be used to reduce foam formation during mixing, thereby lowering the risk of aerosol release at the source.

### Hierarchical configuration of personal protective equipment (PPE)

In this study, quantitative assessments of aerosol exposure concentrations at various stages were conducted, providing data support for implementing precise and efficient protective measures. The study proposed a graded allocation scheme for personal protective equipment (PPE) that aligns with measured risk levels to avoid insufficient or excessive protective measures. The core principle is that during operational phases, the protection level should be positively correlated with the aerosol concentration (PFU/m³). Specific details are presented in Table 7.

**Table 7.** PPE classification configuration scheme.

| Risk Grade | Operate Link | Recommended PPE Configuration | Wearables Requirements | Replacement Frequency |
| --- | --- | --- | --- | --- |
| High Risk | Sample homogenization | N95 mask (GB2626-2019) + anti-fog goggles (GB14866-2022) + body protection suit (GB19082-2009) + double-layer nitrile gloves (thickness ≥ 0.1 mm) + anti-slip shoe cover | The mask fits snugly against the face with no air leakage; goggles cover the entire eye area for full protection; zip up the protective suit to the top and tighten cuffs and ankles to prevent aerosol entry; double-layer nitrile gloves fit snugly on hands without damage. | At the end of each batch, replace all PPE to avoid cross-contamination |

| **Risk Grade** | **Operate Link** | **Recommended PPE Configuration** | **Wearables Requirements** | **Replacement Frequency** |
| --- | --- | --- | --- | --- |
| **High Risk** | Nucleic acid extraction | N95 mask + protective mask (covering the nose and mouth and jaw) + isolation gown (GB/T32610-2016) + single layer nitrile gloves | The protective face mask is seamlessly connected to the mask to ensure full protection of the face; the isolation sleeve is inserted into the glove to prevent aerosol from entering through the cuff. | Gloves are changed every 10 samples; isolation gowns are changed after each batch of experiments |
| **Low Risk** | Sample collection | Medical surgical mask (YY0469-2011) + disposable isolation gown + single-layer nitrile gloves | The mask ear hook is firmly fixed to cover the mouth and nose to ensure respiratory protection; the disposable isolation clothing is worn neatly without damage. | Gloves are changed every 5 samples collected; isolation gowns are changed after daily experiments |
| **Low Risk** | PCR amplification and result analysis | Medical surgical mask + disposable hat + single-layer nitrile glove | The disposable cap covers all hair to prevent hair loss and contamination of the experimental environment; the gloves are intact and fit the hands. | In the PCR amplification step, gloves should be changed every time the reaction tube batch is replaced; in the result analysis step, there is no special requirement, and gloves can be replaced timely according to the actual situation. |

In addition, a standardized personal protective equipment (PPE) removal procedure was established on the basis of the principle of "pollution first, then cleaning and disinfection": (1) immediately remove the outer layer of gloves and disinfect hands with 75% ethanol; (2) when removing protective suits/isolation gowns, roll them up from the inside to avoid contact with contaminated surfaces; (3) when removing goggles/masks, grip the shoulder straps firmly to prevent lens contact; (4) when removing masks, secure the ear hooks to prevent external contamination; and (5) after removing the inner layer of gloves, disinfect hands with 75% ethanol and rinse them under running water according to the seven-step handwashing procedure for at least 20 s.

### Laboratory environmental control and aerosol monitoring system

On the basis of research findings regarding aerosol spatial distribution characteristics, concentration levels, and influencing factors, a comprehensive environmental safety system comprising three components—"regional control, active monitoring, and emergency response"—should be established from the perspectives of engineering control and management processes. (1) Spatial Zoning and Ventilation Optimization: In terms of spatial zoning, PCR laboratories are classified into "high-risk areas" (including sample homogenization and nucleic acid extraction zones) and "low-risk areas" (amplification and analysis zones) on the basis of risk levels. A 1.5-m-wide buffer corridor equipped with air curtains (wind speed ≥8 m/s) is established between these areas to prevent aerosol transmission. To optimize ventilation systems, high-risk areas employ independent ventilation systems, increasing air exchange rates to 15–20 times per hour (in contrast to 6–12 times per h in conventional PCR laboratories), thereby accelerating air renewal to dilute and disperse aerosols. Exhaust outlets are fitted with HEPA filters with an efficiency ≥99.97%, which are replaced every six months and subjected to integrity testing. Biosafety cabinet maintenance requires 30 min of ventilation prior to daily use to stabilize airflow, followed by continued operation for 30 min post-use to disperse aerosols. The internal walls and work surfaces are wiped weekly with a 500 mg/L chlorine disinfectant solution, and the airflow velocity is monitored monthly to ensure compliance with the standards. (2) Aerosol Monitoring and Early Warning Mechanism: In terms of monitoring frequency, high-risk areas (sample processing zones) undergo monthly monitoring, while low-risk areas (amplification and analysis zones) are monitored quarterly. When handling samples with high viral loads (severe infection patient samples with Ct values <25), the monitoring frequency is increased to biweekly. This dual-monitoring approach integrates the ΦX174 phage simulation technology developed in this study with qPCR for actual viral detection. If either monitoring method detects aerosol concentrations ≥10³ PFU/m³ (or ≥10³ copies/m³), the system will immediately trigger an alarm.

The emergency response protocol is as follows: (1) the experimental operations should be suspended, and personnel should be evacuated; (2) the ventilation system should be shut off to prevent aerosol transmission; (3) spray disinfectant (500 mg/l chlorine-containing solution, concentration of 20 ml/m³) should be applied for 30 min; and (4) ventilation should be resumed after disinfection and remonitoring should be conducted after 24 h. Experimental activities may be resumed only when the concentration falls below the threshold (10³ PFU/m³ or <10³ copies/m³).

## Conclusions

In this study, the ΦX174 bacteriophage was used as an indicator virus to quantify aerosol concentrations in BSL-2 PCR laboratory workflows. The results demonstrated that sample homogenization (5.67±1.23)×10⁴ PFU/m³ was classified as an extremely high-risk procedure and that nucleic acid extraction (3.45±0.89)×10⁴ PFU/m³ was classified as a high-risk step, whereas sample collection, PCR amplification, and result analysis were categorized as low-risk operations. Significant differences in aerosol concentrations were observed across stages (F = 156.32, P < 0.001). The viral load in the samples strongly positively correlated with the aerosol concentration (r = 0.926; P < 0.001), with high-risk aerosol concentrations exhibiting linear attenuation as the sampling distance increased. This study confirmed that the biological characteristics of the ΦX174 bacteriophage closely matched those of pathogenic viruses, with a detection sensitivity of 10¹ PFU/ml, indicating its suitability for BSL-2 laboratory environments as an ideal indicator viral vector. The proposed strategies, including high-risk equipment upgrades, graded personal protective equipment allocation, and environmental monitoring and early warning mechanisms, effectively reduced aerosol exposure risks in PCR laboratories, providing scientific evidence and standardized operational protocols for laboratory occupational health management.

## Data Availability

The data underlying this article will be shared on reasonable request to the corresponding author.

## Acknowledgments

We thank the Hubei Province Key Laboratory of Occupational Hazard Identification and Control, Wuhan University of Science and Technology, for technical support.

